# Correlation of IL-6, Coping, &Depression in Adolescent and Adult Patients Receiving Hemodialysis: A Comparative Study

**DOI:** 10.1101/2022.05.31.22275844

**Authors:** Sanaa A. Kamal, Aref A. Khoweiled, Maged E. Gomaa, Sameh A. Al-Dawy, Reham M. Kamel, Doaa R. Ayoub

**Author notes:** **Corresponding Author:** Doaa R. Ayoub, MBBCH, MSc, MD, Faculty of Medicine, Cairo University, Kasr Alainy Street, P.O. box: 11562, Cairo, Egypt). **Sanaa A. Kamal**, Professor of Psychiatry, Faculty of Medicine - Cairo University, Cairo- Egypt. **Aref Khoweiled**, Professor of Psychiatry, Department of Psychiatry, Faculty of Medicine, Cairo University, Cairo- Egypt. **Maged E. Gomaa**, Associate Professor of Psychiatry, Faculty of Medicine - Cairo University, Cairo- Egypt. **Sameh A. Al-Dawy**, Lecturer of Internal Medicine, Faculty of Medicine - Cairo University, Cairo- Egypt. **Reham M. Kamel**, Lecturer of Psychiatry, Department of Psychiatry, Faculty of Medicine, Cairo University, Cairo- Egypt. **Doaa R. Ayoub**, Lecturer of Psychiatry, Department of Psychiatry, Faculty of Medicine, Cairo University, Cairo- Egypt.

## Abstract

**Background:** The research has long reported that depression and stress are highly prevalent among patients with end-stage kidney disease requiring dialysis. Growing studies suggest that the inflammatory gene IL-6, in particular, contributes to the etiology of depression & stress by affecting the function of serotonin; one of the other explanations is by causing hypercortisolemia with overstimulation of the HPA axis. This study aimed to assess the inflammatory gene IL-6 in adolescent & adult patients with end-stage kidney disease receiving hemodialysis. Furthermore, to assess the coping skills of those patients upon the emergence of depressive symptoms & stress levels.

**Method:** One hundred and twenty-one patients receiving hemodialysis were recruited in a cross-sectional study from King Fahd Unit at Kasr Al-Ainy Hospital, Cairo University, Egypt. They were assessed for depression using Beck’s II depression inventory, stress by the stress perceived scale, coping by the short COPE questionnaire & they had inflammatory gene IL-6 level measured.

**Results:** Seventy-six patients showed depressive symptoms; adults were more depressed than adolescent patients, yet adolescents were subjected to more stress. The mean level of Interleukin-6 was 148.0±50.5pg/ml, which is higher than average. Perceived Stress Scale mean scores showed a statistically significant difference between depressed adults and adolescents (*p*=0.050).

**Conclusions:** Inflammatory gene IL-6 shows a higher level in depressed adolescent and adult patients receiving hemodialysis. Adult patients are more depressed than adolescent patients with end-stage kidney disease yet coping strategies are far better in depressed adolescents than adults.

## BACKGROUND

WHO statistics for Egypt showed that End-Stage kidney Disease has shown almost exponential growth in the last years ^**(1)**^.

It has been reported in the literature that depression is highly prevalent among patients with End-stage renal disease requiring dialysis ^**(2)**^.

Growing studies suggest that inflammatory cytokines especially IL-6, in particular, contribute to the etiology of depression by affecting the function of serotonin, one of the other explanations is by causing hypercortisolemia with overstimulation of the HPA axis resulting in depression ^**(3)**^.

Several researchers support the supposition that pro-inflammatory cytokines are involved with depression in renal patients. In particular, there is evidence that depression is associated with *IL-1, IL-6*, tumor necrosis factor-alpha (*TNF-*α), and C-reactive protein (*CRP*) in both the general and ESRD populations ^**(4)**^. Increased cytokine levels are noticed in uremia ^**(5)**^.

Central to most contemporary theories of depression is the notion that stress can initiate cognitive and possibly biological processes that increase the risk for depression ^**(6,7)**^.

Chronic and life-threatening diseases are among stressful factors in humans ^**(8)**^. The Coping strategies are a collection of cognitive and behavioral personal struggles adopted to interpret, comment, and modify stressful situations and result in the suffering relief of these situations.

The main strategies are:-

a. ***Emotion-focused strategies:*** including all attempts to regulate emotional outcomes of stressful events and make an emotional balance through emotional control ^**(9)**^.
b. ***Problem-focused coping strategies:*** that include self-constructive behavior with stressful situations and trying to detect or change the source of stress ^**(10)**^.

Although both strategies bring about a stress reduction, research showed that those individuals who use emotion-focused coping strategies have lower psycho-cognitive adaptation**; w**hile problem-focused coping strategies are seen among those with appropriate adaptation ^**(11)**^. **Curtin et al**. ^**(12)**^ Provided insights into “the transitional experience” of survivors of long-term dialysis. The restructuring of illness beliefs and modification of “illness” cognitions result in positive changes; patients can become “active self-managers of their disease, its treatment, and its manifestations.” “The concept of transition is often advanced as the embodiment of successful adjustment to a chronic illness”. A sense of control over illness influences coping and adjustment to long-term physical health problems ^**(13)**^.

The objective of this study is to assess the inflammatory gene IL-6 in adolescent & adult patients with end-stage kidney disease receiving hemodialysis. And also to assess the coping skills of those patients on the emergence of depressive symptoms.

## METHODS

### Study Characteristics

A convenient cross-sectional sample of one hundred and twenty-one subjects was recruited from the King Fahd Unit of dialysis at Kasr Al-Ainy Hospital, Cairo University over one year.

All procedures performed in this study were in concordance with the ethical standards of our institution - The scientific and Ethical committee of the Psychiatry Department, Faculty of Medicine-Cairo University and with the 1964 Helsinki declaration and its later amendments. Informed written assent from each participant and informed written consent from their parents; were obtained from all individual participants included in the study, and the protocol of assessment was approved by the committee of ethics of The Faculty of Medicine.

### Sample Selection

Patients were recruited after completion of the psychiatric assessment and tools, including the mini-mental state examination.

#### Inclusion criteria

End-Stage Renal Disease (ESRD) patients (age 15 to 60 years), and on chronic hemodialysis.

#### Exclusion criteria

Patients with acute sepsis, chronic infections, immune disease, malignancies, on steroids or immune-modulatory therapies, intellectual disabilities, or major cognitive problems that cannot complete the questionnaires or with end-organ failure other than renal failure. Refusal to consent or participate was also an exclusion criterion.

### Data Collection Tools and Procedure

Patients were recruited from King Fahd Unit after being assessed by the assigned nephrologist. Patients were submitted to psychiatric, medical, and, IL-6 level assessments.

The Psychiatric evaluation included SCID interview applying DSM-IV criteria for depression, perceived stress score (PSS), short COPE, and Short form health survey-36 (SF-36). The medical assessment was conducted to examine recruited subjects and to declare their free state of sepsis or other exclusion criteria and withdrawal of serum levels of interleukin-6 (IL-6) from participating subjects.

Psychiatric assessment was performed through:-

a. *Beck depression inventory:* This is a self-report rating scale measuring the emotional, cognitive, and motivational symptoms of depression ^**(14,15)**^.
b. *The perceived stress scale (PSS-10):* The Arabic version of PSS-10 was found to have adequate reliability and validity. The PSS-10 covers two-factors. The first factor included questions reflecting negative feelings (being upset, angry, or nervous) and inability to handle stress while the second factor included questions expressing positive emotions and ability to act in stressful situations ^**(16.17)**^
c. *Brief (COPE) scale:* The Brief COPE is a self-rating scale that was designed to assess a broad range of coping responses among adolescents and adults for all diseases ^**(18.19)**^ In total, 14 dimensions are covered by this scale (Self-distraction, active coping, denial, substance use, use of emotional support, use of instrumental support, behavioral disengagement, venting, positive reframing, planning, humor, acceptance, religion, and self-blame).
d. *The Short Form (36) Health Survey (SF-36):* The SF-36 is a measure of health status and an abbreviated variant of it, the SF-36, is commonly used in health economics to determine the cost-effectiveness of health treatment ^**(20,21)**^.
e. *SCID I:* is a Structured Clinical Interview for DSM-IV ^**22**^ axis I disorders diagnoses^**(23,24)**^ Those who agreed to sign the informed consent were screened for the presence of depressive symptoms using Beck’s Depression inventory II, patients were grouped into two groups: The depressed group with (BDI-II score ≥ 14) and the Non-depressed group with (BDI-II score < 14).
f. The analysis for inflammatory gene IL-6 was performed at the Department of Bio-Chemistry Faculty of Medicine – Cairo University. Blood was drawn on a serum separator tube (SST) by a trained nurse. Within 30 minutes of collection, centrifugation was done for 15 minutes. Then serum was extracted and stored at ≤-20ºC, Serum IL-6 concentration was measured by using ELISA with a sensitivity of 2 pg/m; using a commercially available human IL-6 ELISA kit (Gen-Probe, Diaclone France).

### Data analysis

All data were computed and analyzed, using SPSS ^**(25)**^ (Statistical Package for Social Sciences - version 16) IBM Corp USA software for statistical analysis. Descriptive statistics were used for illustrating the mean and standard deviation of quantitative data. Qualitative data were presented as percentages and proportions. The variables were analyzed using measures for comparison (T-test, ANOVA, and Chi-square) and measures of association (Pearson correlation and regression). P-value was considered significant if ≤ 0.05.

## RESULTS

Males constituted 52.1% of the study population (63 males). The study included 21 adolescents (age ≤ 21 years old) (17.4%) For adolescents, the average age was 19.9±1.5 and they included 8 males (38.0%). For adults, the average age was 45.1±13.4 and they included 55 males (55.0%). There was no significant difference between adolescents and adults in their gender distribution (p= 0. 102).

As for the clinical assessment of the study subjects, depression was found in approximately 63% of the sample n. 76 (Table 1). The mean score of the perceived stress scale was 18.3±3.7.

**Table (1):** Prevalence of depression using Beck’s Depression Inventory.

|  |  |  |
| --- | --- | --- |
| Beck's Depression Inventory | Total score(n=121) | 16.3±8.3 |
|  | Mild depression( n=29) | 17.7±1.5 |
|  | Moderate depression (n=43) | 23.3±2.7 |
|  | Severe depression(n=4) | 34.5±3.0 |

Depressed dialysis patients had statistically significant lower scores (*p*< 0.05), regarding their venting, planning, and humor scores. Also, dialysis depressed patients showed a trend toward lower active coping and religion (Table 2).

**Table (2).** Comparing coping strategies among depressed and non-depressed dialysis patients using Brief COPE Scale mean scores.

| Brief COPE score | No depression<br>BDI<14<br>(n=45) | Depression<br>BDI>14<br>(n=76) | p-value |
| --- | --- | --- | --- |
| Self-distraction | 5.6±2.4 | 5.0±2.2 | 0.178 |
| Active coping | 5.5±2.6 | 4.8±1.6 | 0.097 |
| Denial | 4.8±2.0 | 4.6±2.0 | 0.710 |
| Substance use | 0.9±1.8 | 0.8±1.9 | 0.737 |
| Emotional support | 5.5±2.2 | 5.6±2.0 | 0.765 |
| Instrumental support | 5.7±2.5 | 5.2±2.1 | 0.222 |
| Behavioral disengagement | 5.0±1.8 | 4.5±1.7 | 0.159 |
| Venting | 5.1±1.7 | 4.4±1.2 | 0.022* |
| Positive reframing | 4.8±2.2 | 5.2±6.0 | 0.630 |
| Planning | 5.6±2.2 | 4.6±1.7 | 0.007* |
| Humor | 5.2±1.8 | 4.5±1.6 | 0.025* |
| Acceptance | 5.2±2.0 | 4.8±1.9 | 0.353 |
| Religion | 5.4±2.2 | 4.7±1.9 | 0.053 |
| Self-blame | 4.4±2.1 | 4.6±1.7 | 0.640 |

Table (3) results showed no statistically significant difference for Beck’s Depression Inventory mean score (*p*= 0. 109) between depressed adults and adolescents, however, Perceived Stress Scale mean scores showed a statistically significant difference between depressed adults and adolescents (*p*=0.050).

**Table (3):** Comparison of mean scores of Beck’s Depression Inventory & Perceived Stress Scale between adult and adolescent depressed dialysis patients.

|  | Depressed Adults (n=62) | Depressed Adolescents (n=14) | *p*-value |
| --- | --- | --- | --- |
| Beck's Depression Inventory | 22.1±4.9 | 19.9±3.3 | 0.109 |
| Perceived Stress Scale | 18.9±3.1 | 20.6±2.7 | 0.050 |

Depressed adolescents showed better all-over coping skills such as self-distraction, venting, religion, emotional support, and instrumental support humor scores but it wasn’t statistically significant (Table 4).

**Table (4).**
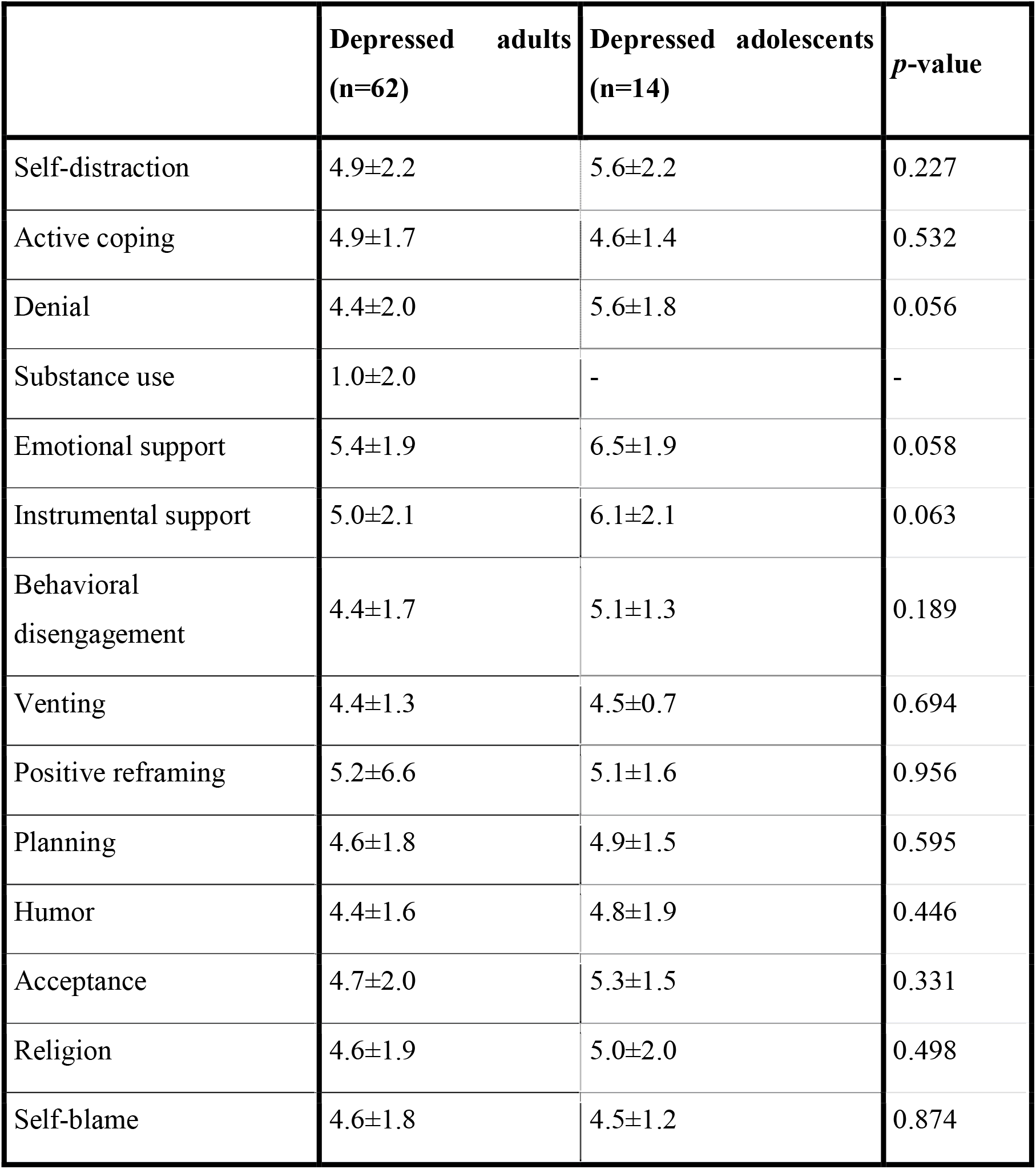
Comparison of coping strategies among adult and adolescent depressed dialysis patients in the study sample using Brief COPE Scale mean scores.

Table (5) showed that the comparison of the mean level of IL-6 was higher than the normal reference range between different groups. Our research showed that the mean level of Interleukin-6 was 148.0±50.5pg/ml. which was increased 30 folds as it was referenced by Mayo Clinic’s Laboratories (Normal reference range ≤ 5 pg/mL) yet it wasn’t statistically significant.

**Table (5):**
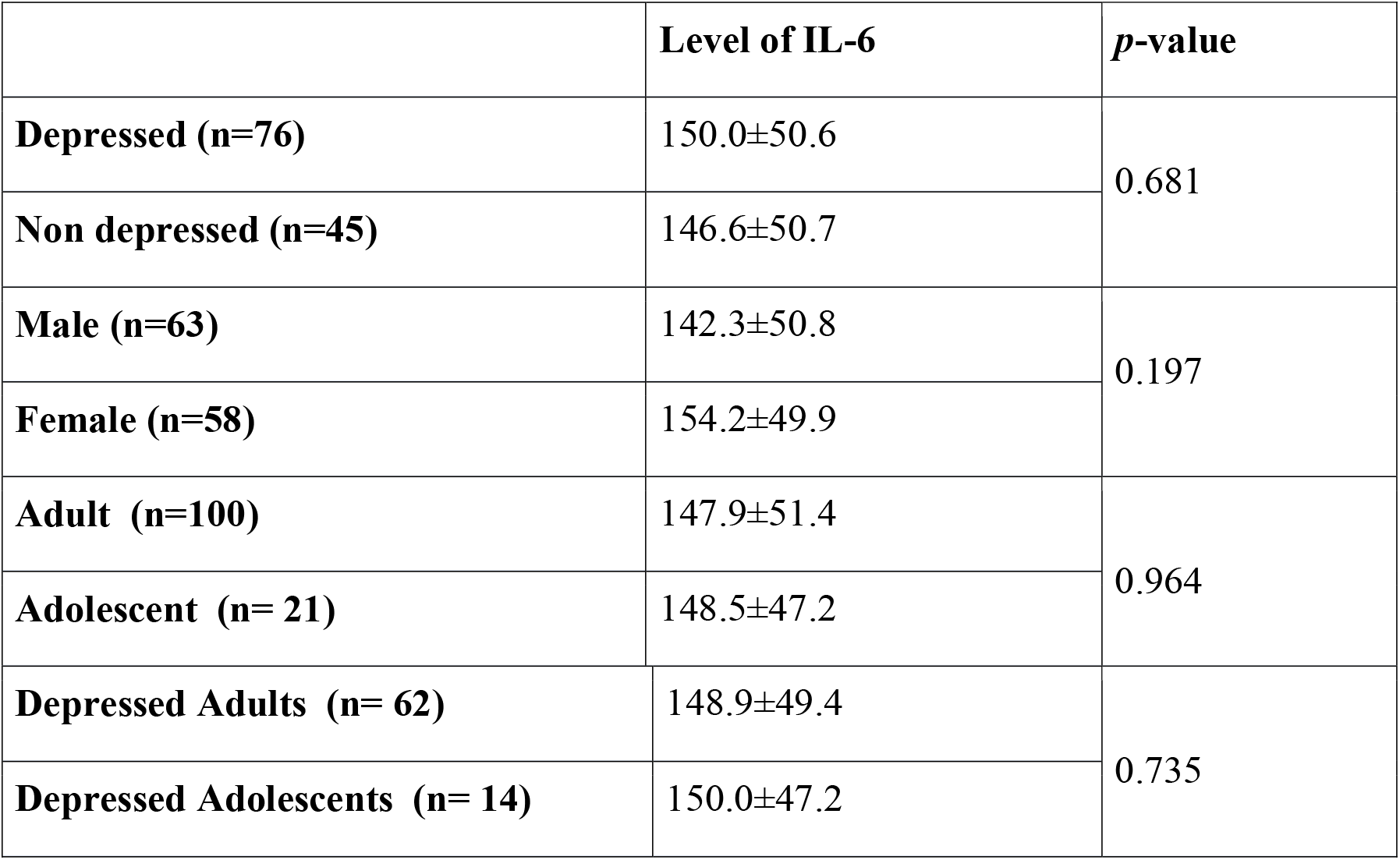
Comparison of Il-6 value between depressed/non-depressed, male/female, and adult/adolescents.

The current study showed that the mean of Interleukin-6 was 148.0±50.5pg/ml. yet there was no statistically significant difference in interleukin-6 between depressed and non-depressed patients, between males and females, and between depressed adults and adolescents. In addition, Interleukin-6 did not correlate with Beck’s Depression Inventory, Perceived Stress Scale, SF-36 scores, or with the duration of illness in the study population (Table 6).

**Table (6):** Correlations between IL-6 value and (BDI, PSS, SF-36) scores and duration of dialysis.

| **Interleukin-6** | **Correlation coefficient** | ***p*-value** |
| --- | --- | --- |
| Beck's Depression Inventory | -.052 | 0.574 |
| Perceived stress score | -.057 | 0.534 |
| SF-36 | .029 | 0.755 |
| Duration of illness | -0.100 | 0.277 |

Graph (1) shows that the higher the duration of illness the higher Beck’s Depression Inventory scores (r-value .358, P-value <.001).

**Graph 1:**
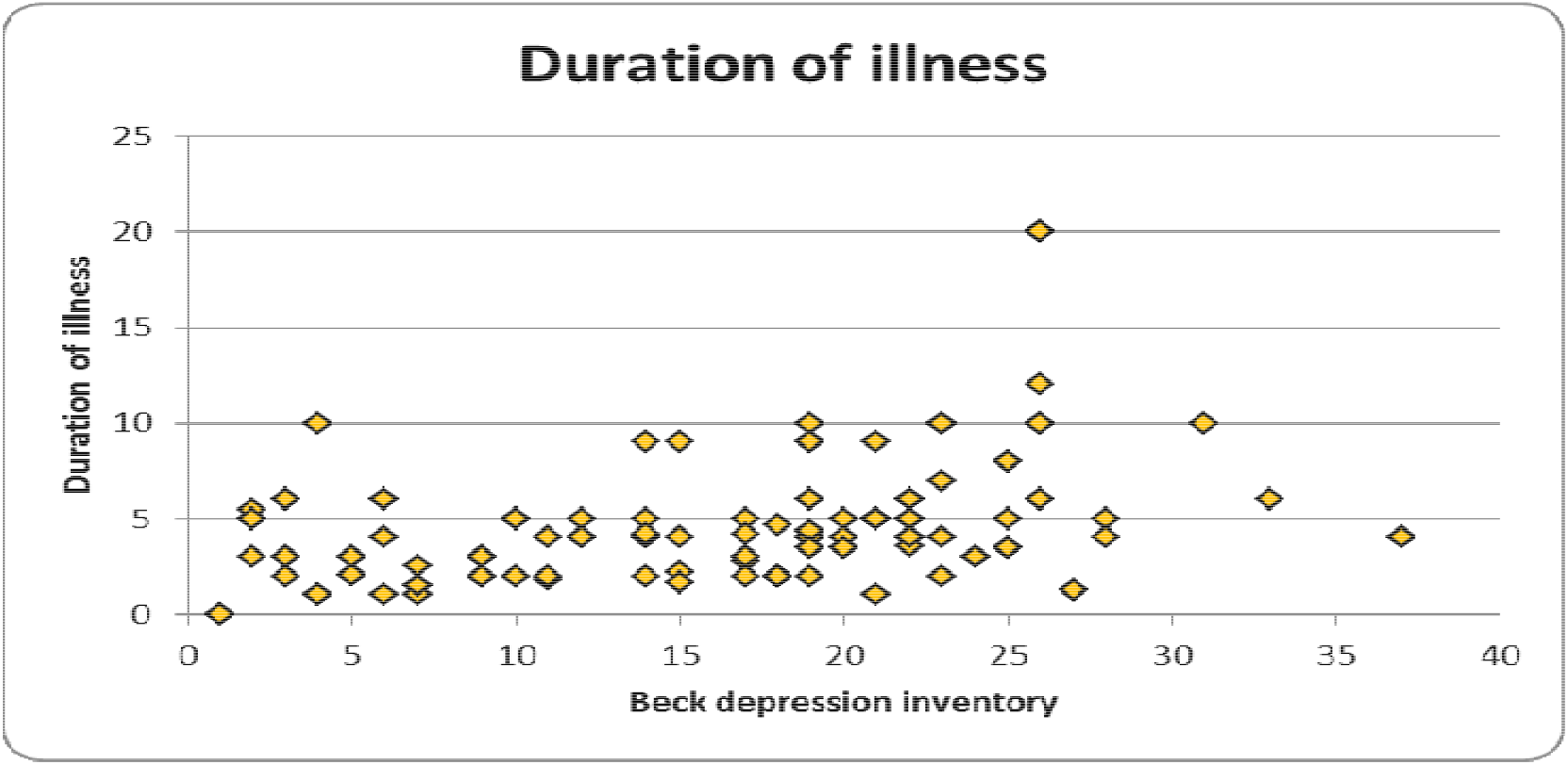
Correlation between Beck’s Depression Inventory results and duration of Dialysis.

## DISCUSSION

This was a cross-sectional study that recruited 121 dialysis patients to assess the inflammatory gene IL-6 in depressed patients with kidney disease, and their coping and stress levels. The study included 21 adolescents and 100 adults. The discrepancy in the number of both groups is related to the relative decrease in the prevalence of ESRD in the teenage group. This was consistent with the Egyptian renal registry that the prevalence of Chronic Kidney Disease (CKD) in adults was 10%, while in adolescents was 2.5% ^**(26)**^. As regards the prevalence of depression, 69 patients were diagnosed with depression using SCID I, and 76 patients were diagnosed with depression using Beck’s Depression Inventory (a cut-off score ≥14). This prevalence was in line with the literature, which rates depression in ESRD as between 10 to 100% ^**(27,28,29)**^. Variations in prevalence rates of depression in different studies may be related to the differences in the used tool and the cut-off of the used scale.

Recently, many authors have tried to find a connection between depression and aggravated inflammatory status of patients with ESRD especially IL-1, IL-6, and, IL-10 ^**(30)**^.

Our research showed that the mean level of Interleukin-6 was 148.0±50.5pg/ml. which was increased 30 folds as it was referenced by Mayo Clinic’s Laboratories (Normal reference range ≤ 5 pg/mL) which was consistent with several studies ^**(31) (32) (33)**^ that reported increased levels of IL-6 in HD patients.

Several researchers ^**(34)(35)(36)**^ concluded that higher levels of IL-6 in HD patients might be correlated with the severity of depression. On the contrary, in our study, there was no statistically significant difference in interleukin-6 between depressed and non-depressed patients, between males and females, and between depressed adults and adolescents. In addition, the Interleukin-6 level did not correlate with Beck’s Depression Inventory, Perceived Stress Scale, SF-36 scores, or the duration of illness in the study population. This was in line with **Hung et al**., ^**(37)**^ who did not find any relationship between BDI score and the level of IL-6 neither in the whole group of patients with ESRD nor in the group of depressed patients on the HD. They explained the difference in their results to the literature that HD patients recruited in their sample were exposed to several cytokine– inducing factors, such as contact with dialysis membranes, contamination by endotoxin, and non-endotoxin cytokine-inducing factors which can disturb their relationship. The dialysis membrane especially the used Curephane membranes can stimulate monocytes, either directly or through activation of the complement (increasing cytokine production) which is another specific inflammatory factor in HD patients ^**(38)**^.

As regards the elevation of inflammatory gene IL-6 in the adolescent group, despite the difference in etiologies (congenital disease); This provided more evidence for the explanation that the used membranes and dialysate played a major role in the elevation of cytokine levels in this study sample, which masked the effect of IL-6 relation in depression.

Adults tended to show higher scores on Beck’s Depression Inventory than adolescents. This was consistent with **Chiang et al**. ^**(39)**^ study who found that depression tended to increase with age. However, Perceived Stress Scale showed a statistically significant difference between depressed adults and adolescents (*p*=0.050), being higher in adolescents than adults.

Results revealed that adolescents were more prone to stress than adults, this might be attributed to the lack of sense of autonomy at this age and the poor body image due to dialysis fistulas, stunted growth, and earthy look which decreased their self-esteem. Although adolescents were more prone to stress they showed lower scores on depression than adults. This could be explained that adults had an increased duration of illness which increases their burden due to their responsibilities the chronicity of illness and persistent nature of disease prohibits them from committing it. Moreover, most of the donations offered to transplantation operations in the King Fahd Unit are directed to those at a young age, which increases their hope for a better life.

On the other side, adolescents with chronic kidney disease reported more family involvement, better safety and health practices, and better social problem-solving skills ^**(40)**^. It was noticed during the study that adolescents attended their dialysis sessions, accompanied by their relatives while most of the adult patients attended their sessions alone.

Coping with ESRD is critical, as it influences the patients’ ability to manage their lives and adjust to life demands ^**(41)**^. In this study, depressed patients compared to non-depressed, had significantly lower scores, regarding their venting, planning, and humor scores. In addition, depressed patients showed a trend towards lower active coping. It was shown that the most used strategies with depressed patients were emotional and instrumental support. This was in line with studies that showed individuals who use emotion-focused coping strategies have lower psycho-cognitive adaptation ^**(42)**^. This was in line with other research ^(11)(43)^ which found that patients on hemodialysis tend more to use emotion-focused coping strategies rather than problem-solving strategies.

## Strengths & limitations

To the knowledge of the researchers, this was the first study to address the correlation of interleukins to depression & stress in hemodialysis patients in Egypt and to conduct a comparison between adult and adolescent patients, especially regarding the biological etiology of depression, the large study sample, and the chance for recruiting patients from King Fahd Unit which is considered one of the largest catchment areas for patients all over Egypt; are all considered strengths.

As for limitations, the study at hand didn’t assess other inflammatory cytokines (IL-1, IL-10, and, TNF) and their relation to depression due to the limited financial resources. Patients on peritoneal dialysis which causes less inflammatory reactions were not assessed, as peritoneal dialysis was not available at Kasr Al-Ainy Hospital during the study period. Comparison to patients using dialysis membranes other than Curephane membranes was not done; nor was the assessment of IL-6 level after the dialysis session.

## Conclusions

Serum IL-6 shows a higher level in depressed adolescent and adult patients with end-stage kidney disease receiving hemodialysis. Adult patients are more depressed than adolescent patients on hemodialysis with ESRD, coping mechanisms are superior in depressed adolescents compared to adults.

## Data Availability

All data produced in the present study are available upon reasonable request to the authors

## Abbreviation

CRP: C-Reactive Protein
DSM-IV: Diagnostic and Statistical Manual of Mental Disorders 4^th^ Edition
ESRD: End-Stage Renal Disease
HD: Haemodyalyisis
IL-6: Interleukin 6
PSS: Perceived Stress Scale
SCID I: Structured Clinical Interview for DSM-IV
SF-36: Short Form Health Survey-36
SST: Serum Separator Tube
TNF-α: Tumor Necrosis Factor-Alpha

## Declarations

### Ethics approval and consent to participate

All procedures performed in this study were in concordance with the ethical standards of our institution - Scientific and Ethical committees of the Psychiatry Department, Faculty of Medicine-Cairo University and with the 1964 Helsinki declaration and its later amendments. At the time of conducting this study, the reference number was not applicable as the ethical approval was only a necessity at the departmental level without any other prerequisites from the University.

Informed written assent from each participant and informed written consent from their parents were obtained from all individual participants included in the study,

### Consent for publication

Not Applicable

### Availability of Data and Materials

The data used and analyzed during the current study are available from the corresponding author on reasonable request.

### Competing interests

On behalf of all authors, the corresponding author states that there is no conflict of interest and that the study received no funds and was funded by the researchers.

### Funding

The research was funded by the researchers

### Authors’ contributions

All authors had made a substantial contribution to the design of the work, data collection, & interpretation, writing the manuscript, revising it, and approving the final version.

## Notes

### Competing Interest Statement

The authors have declared no competing interest.

### Funding Statement

This study did not receive any funding

### Author Declarations

All procedures performed in this study were in concordance with the ethical standards of our institution - The scientific and Ethical committee of the Psychiatry Department, Faculty of Medicine- Cairo University and with the 1964 Helsinki declaration and its later amendments. Informed written assent from each participant and informed written consent from their parents; were obtained from all individual participants included in the study, and the protocol of assessment was approved by the committee of ethics of The Faculty of Medicine.

